# Factors associated with breast lesions among women attending select teaching and referral health facilities in Kenya: a cross-sectional study

**DOI:** 10.1101/2024.08.08.24311692

**Authors:** Josephine Nyabeta Rioki, Marshal Mweu, Emily Rogena, Elijah M. Songok, Joseph Mwangi, Lucy Muchiri

## Abstract

**Background:** Despite extensive research on the risk factors for breast cancer, little is known about the factors contributing to other breast lesions, of which some may indicate an increased risk for this disease. The objective of this study was to identify the risk factors for breast lesions among women with breast lumps seeking care at select teaching and referral hospitals in Kenya between December 2016 to December 2019.

**Methods:** An analytical cross-sectional study design was used to investigate the risk factors for breast lesions among 651 women with breast lumps. Data was collected using a semi-structured questionnaire. A multivariable logistic regression model was used to evaluate the impact of the predictors on the breast lesions. Ethical approval was granted by Kenyatta National Hospital (KNH) and the University of Nairobi Ethics and Research Committee (UoN-ERC) under study number P334/04/2016.

**Results:** The median age of participants was 30 years (range 16-87), with the majority having secondary education and belonging to the Kikuyu ethnic group. Parity, exercise, and contraceptive use were significant predictors of breast lesions identified. Nulliparous women had significantly lower odds of developing malignant (aOR: 0.11; 95% CI: 0.05-0.26), and suspicious (aOR: 0.23; 95% CI: 0.07-0.71) lesions. Regular exercise was associated with lower odds of both malignant and suspicious lesions. Conversely, contraceptive use increased the odds of developing atypical (aOR 0.92; 95% CI: 0.28-2.98) lesions, suspicious (aOR 0.33; 95% CI: 0.14-0.78), and malignant lesions (aOR of 0.31; 95% CI: 0.18-0.55).

**Conclusion:** Exercise, parity, and hormonal contraception were independently found to be significant predictors of breast lesions. These findings underscore the necessity for personalized risk reduction strategies and emphasize the importance of understanding the risk factors for both benign and malignant breast disease to inform public health policies.

## Background

The most frequently diagnosed breast disease among women worldwide is breast cancer (BC), also referred to as a malignant breast lesion. About 80% of patients with the disease are individuals aged >50 [1]. It is recorded that BC incidence rates are higher but with low mortality rates in transitioned versus transitioning countries [2]. The incidence is noted to be higher among white women unlike black women, who have higher mortality [3]. In Kenya, the rise in BC incidence has been reported with a prediction that by 2025, it will surge by a devastating 35% [4].

About half of breast cancers can be explained by known risk factors (such as age and female gender) while the other half may not be known [5]. However, even with known risk factors, it is not certain that females with other risk factors will get BC. Based on calculated incidence rates, about 12.9% of women will develop BC in their lifetime. Risk factors for BC are classified into modifiable and non-modifiable factors. Modifiable risk factors include education level, parity, breastfeeding, passive smoking, and obesity. Non-modifiable risk factors include age, age at menarche, menopause, family history of any type of cancer, and family history of BC [6-8].

Global studies have examined the impact of reproductive factors on BC risk, including age at menarche, age at first childbirth, age at menopause, parity, breastfeeding, number of pregnancies, and the occurrence of abortion [9, 10]. Existing literature indicates that changes in reproductive patterns, such as lower parity, delayed pregnancies, and shorter breastfeeding duration, contribute to an increased susceptibility to BC in women [11]. Another previous research has also suggested that prolonged exposure to endogenous oestrogen due to early menarche, delayed first childbirth, late menopause, or exogenous exposure through hormone replacement therapy or oral contraceptive use, are associated with a higher risk of BC [10]. Physical exercise has been shown to affect the fourteen hallmarks of breast cancer, thereby preventing the development of the disease [12].

Though extensive research has been done focusing on the factors associated with BC, little is documented on the factors associated with other breast lesions such as benign, atypical, and suspicious lesions. Some of these lesions are associated with the risk of developing the disease. Notably, among women diagnosed with invasive breast cancer, 30% had benign breast disease previously [13]. Among all atypical breast lesions, atypical ductal hyperplasia (ADH) has the highest likelihood of progressing to malignancy [14].

Since there are differences in population structure, cultural factors, and the adoption of Western lifestyles across different regions, this study aimed to identify the factors associated with BC and other breast lesions among women in Kenya. Understanding these context-specific factors is crucial to inform the development of personalized risk reduction strategies and public health Policies. We report the association between parity, hormonal contraception, and physical exercise with breast lesions from a hospital-based cross-sectional study of 651 women in Kenya.

## Material and methods

### Ethical considerations

Approval to conduct this study was granted by the Kenyatta National Hospital and the University of Nairobi Ethics and Research Committee (study no. P334/04/2016) as well as the institutional review boards of the select facilities. All participants provided written informed consent. The study was conducted following the Helsinki Declaration.

### Study design

This was a cross-sectional study aimed at investigating the factors associated with breast lesions (benign, atypical, suspicious, and malignant breast lesions) among women attending select teaching and referral health facilities in Kenya between December 2016 to December 2019. The study was reported as per the STROBE guidelines for reporting observational studies [15]. This work is part of a larger study, [reference number P334/04/2016] whose main objective was to characterize breast cancer phenotypes and genotypes among women diagnosed with breast cancer at selected teaching and referral health facilities in Kenya.

### Study sites

This study was conducted in two major health facilities in Kenya. The sites were selected due to their status as major referral centres with well-established and busy fine needle aspiration (FNA) clinics that attend to a significant number of women with breast lumps.

Kenyatta National Hospital [KNH] is a Level 6 teaching and national tertiary referral health facility situated in the capital city. It is also a teaching hospital for the University of Nairobi Faculty of Health Sciences. The hospital has FNA clinic that runs once a week and receives about 10-15 women with breast lumps a week.

Nakuru County Referral Hospital [NCRH] is a large county referral hospital located in Nakuru town west of Nairobi. The hospital has FNA clinic that runs once a week. It receives about 10 women with breast lumps per week.

### Study population, eligibility, and selection of participants

The study population consisted of all women with palpable breast lumps presenting to the FNA clinics at NCRH and KNH for care. To be eligible for participation in the study, a woman had to meet the criteria which included the presence of a palpable breast lump and the provision of written informed consent for participation. Women with breast lumps due to trauma were excluded from the study.

### Outcome definitions

A breast lesion was defined as a mass that could either be classified as benign, atypical, suspicious of malignancy, or malignant (cancerous). The classification was based on the International Academy of Cytology Yokohama System (IAC YS) as follows:

A benign lesion is a diagnosis made in cases with unequivocally benign cytological features, which may or may not be diagnostic of a specific benign lesion. The criteria include a pattern of predominantly large, cohesive monolayered sheets of uniform ductal epithelial cells or cohesive 3-dimensional epithelial tissue fragments showing streaming of epithelial cells around irregular slit-like holes (secondary lumina’); there may be a mix of smaller tissue fragments and sheets, but dispersal is usually not prominent [16].

Atypical lesion is defined as the presence of cytological features seen predominantly in benign processes or lesions, but with the addition of some features that are uncommon in benign lesions, and which may be seen in malignant lesions. The criteria include, high cellularity, dispersal of intact single cells, varying degrees of nuclear enlargement and pleomorphism and the presence of nucleoli, -features seen in both fibroadenoma and carcinomas [16].

Suspicious of malignancy is defined as the presence of some cytomorphological features which are usually found in malignant lesions, but with insufficient malignant features, either in number or quality, to make a definitive diagnosis of malignancy. The criteria include, high cellularity, overlapping of the cytological criteria for benign and malignant tumors, large epithelial tissue fragments, with some showing a cribriform or micropapillary architecture, in association with smaller tissue fragments and plentiful dispersed cells showing low- to intermediate-grade nuclear atypia [16].

A malignant cytopathological diagnosis is an unequivocal statement that the material is malignant. The criteria include the following: for high grade carcinomas, there is predominantly discohesive smaller tissue fragments with some dispersed single cells, moderate to high N:C ratio, marked irregular chromatin and prominent nucleoli. For low grade carcinoma, high cellularity, predominance of large irregular 3D epithelial tissue fragments with some smaller fragments and dispersed single cells [17]. These are features of breast cancer.

### Data collection and study variables

Three research assistants were recruited and trained to assist with data collection. Quantitative data were collected using structured questionnaires. Information gathered included: demographics (Age, ethnicity, education level, employment status), family history of breast cancer, reproductive factors (menstrual cycle, menstrual status, parity), hormonal contraceptive use, and lifestyle factors (history of smoking, alcohol history, and exercise). The predictors assessed in this study are summarised in Table 1.

**Table 1:** Predictor variables and their measurements.

| Variables | Measurements |
| --- | --- |
| Age (continuous) | Captured in years |
| Level of education (ordinal) | Assessed in four levels: None, primary, secondary, and tertiary |
| Ethnicity (nominal) | Major tribes in Kenya captured as specific tribes (Kikuyu, Luo, Luhya, Kalenjin, Kamba) and minor tribes captured as others |
| Employment status (nominal) | Captured as employed or unemployed |
| Menstrual cycle (nominal) | Captured as regular or irregular |
| Menstrual status (nominal) | Captured as premenopausal or post-menopausal |
| Parity (nominal) | Captured as nulliparous or parous |
| Contraception use (nominal) | Captured as those who had ever used either implant, oral and injection forms at any given time, or implant and oral only, injection only, oral only, implant only and never used any method |
| Family history of BC (nominal) | Captured as yes for those with family history or no for those without family history |
| Smoking history (nominal) | Captured as yes for those who have ever smoked or no for those who have never smoked. |
| Alcohol history (nominal) | Captured as yes for those who have ever taken alcohol and no for those who have never taken alcohol |
| Exercises (nominal) | Captured as yes for those who perform regular exercises and no for those that do not |

### Data processing and statistical analysis

Responses for qualitative variables from the questionnaires were coded before data entry. The data were entered in an MS Excel spreadsheet and exported to Stata v13 software for cleaning and analysis. Continuous variables were summarized using the median and range. For categorical variables, proportions were computed. To assess the association between the study predictors and the breast lesion outcome, a univariable multinomial logistic regression model was used. Predictors with p≤0.2 in the univariable analysis were included in the multivariable analysis. Due to small number of observations in some categories of contraceptive use, this variable was recategorized into a binary variable: use versus no use.

For the multivariable analysis, significant variables from the univariable analysis were offered to a multinomial logistic regression model and their significance was evaluated at a strict p≤0.05. To minimize confounding, non-significant variables were only excluded from the model when the coefficients of the remaining variables did not change by more than 30%. The validity of the final parsimonious model was assessed by testing the assumption of independence irrelevant alternatives using the Hausman-McFadden test. The null hypothesis for the test is that the model is valid at p*>*0.05.

## Results

### Descriptive statistics

A total of 651 women with breast lumps were eligible for inclusion in the study. The median age for the participants was 30 years (Range: 16-97 years). Most women had attained secondary education (45.9%, n=299) while 0.6% (n=4) had no formal education. Kikuyus were the majority (54.5%, n=355) with breast lumps followed by other tribes (19.4%, n=126). Most participants had formal employment (54.5%, n=355). The majority (83.3%, n=542) of the participants were premenopausal. Those with regular menstrual cycle were 87% (n=569) and 52.8% (n=344) were parous while 47.2% (n=307) were nulliparous. Among women using hormonal contraception methods, 29.3 % (n=190) had used oral pills only while 0.9% (n=6) had used three methods (oral, injection and implant) in their lifetime. Among the study participants, 10.5% (n=68) were diagnosed with malignant lesions (breast cancer), 4.2% (n=27) with lesions suspicious of malignancy, 1.8% (n=12) atypical cytologic findings, and 83.6% (n=544) benign lesions. The descriptive statistics for the predictors of breast lesions are displayed in Table 2.

**Table 2:** Descriptive statistics of predictors of breast lesions among women with breast lumps attending two select teaching and referral health facilities in Kenya (n=651)

| Variable | Responses | Frequency n (%) |
| --- | --- | --- |
| Age (years) | 16-97 | - |
| Level of education | None | 4(0.6) |
|  | Primary | 167(25.7) |
|  | Secondary | 299(45.9) |
|  | Tertiary | 181(27.8) |
| Ethnicity | Kamba | 48(7.4) |
|  | Kalenjin | 26(4) |
|  | Luhya | 32(4.9) |
|  | Luo | 64(9.8) |
|  | Kikuyu | 355(54.5) |
|  | Others | 126(19.4) |
| Employment status | Yes | 355(54.5) |
|  | No | 296(45.5) |
| Menstrual cycle | Regular | 569(87) |
|  | Irregular | 82(12) |
| Menstrual status | Pre-menopause | 542(83.3) |
|  | Menopause | 109(16.7) |
| Parity | Parous | 344 (52.8) |
|  | Nulliparous | 307(47.2) |
| Contraception use | Injection+Implant only | 4(0.6) |
|  | Oral+Injection+Implant | 6(0.9) |
|  | Oral+Implant only | 13(2) |
|  | Oral+Injection only | 19(2.9) |
|  | Implant only | 26(4) |
|  | Injection only | 65(10) |
|  | Oral only | 190(29.3) |
|  | None | 326(50.2) |
| Family history of BC | Yes | 32(4.9) |
|  | No | 618 (95.1) |
| Smoking history | Yes | 12(1.8) |
|  | No | 639(98.2) |
| Alcohol history | Yes | 40 (6.1) |
|  | No | 611(93.9) |
| Exercise | Yes | 326(50.1) |
|  | No | 325(49.9) |

Key: n; number of respondents, %; percentage

### Logistic regression analyses

#### Univariable analysis

Based on the results from univariable analysis, age, ethnicity, level of education, employment status, menstruation status, parity, contraceptive use, family history of BC, and exercise were significantly associated with breast lesions at p≤0.20 (Table 3). These variables were offered to a multivariate multinomial model to minimize confounding.

**Table 3:** Univariable analysis of the risk factors for breast lesions among women with breast lumps attending two select referral health facilities in Kenya, using a multinomial logistic regression model.

| **Factors** | **Atypical lesions**<br>**cOR (95% C.I)** | **Suspicious lesions**<br>**cOR (95% C.I)** | **Malignant lesions**<br>**cOR (95% C.I)** | **P-value** |
| --- | --- | --- | --- | --- |
| **Age in years\*** | 1.02(0.98, 1.06) | 1.04(1.01, 1.06) | 1.06(1.04, 1.07) | *<0.001* |
| **Ethnicity\*** |  |  |  | *0.149* |
| Kamba | 1.62(0.14, 18.51) | 0.46(0.06, 3.92) | 3.50(1.49, 8.21) |  |
| Kalenjin | 2.36(0.21, 27.22) | 0.68(0.08, 5.77) | 0.73(0.15, 3.45) |  |
| Luhya | 2.00(0.17, 22.91) | 0.57(0.07, 4.85) | 1.23(0.37, 4.09) |  |
| Luo | 1.79(0.25, 13.07) | 5.39x10<sup>-6</sup>(-, -) | 0.55(0.17, 1.77) |  |
| Kikuyu | 0.86(0.16, 4.50) | 0.84(0.34, 2.07) | 0.82(0.41, 1.63) |  |
| Others (Ref) | 1 | 1 | 1 |  |
| **Level of education\*** |  |  |  | *0.008* |
| None | 0.00(0,-) | 0.00(0,-) | 6.22(0.59, 65.92) |  |
| Primary | 6.72(0.78, 58.17) | 6.27(1.76, 22.29) | 3.43(1.54, 7.68) |  |
| Secondary | 4.06(0.48, 34.03) | 2.26(0.61, 8.33) | 2.63(1.23, 5.62) |  |
| Tertiary (Ref) | 1 | 1 | 1 |  |
| **Employment status\*** |  |  |  | *<0.001* |
| Employed | 0.97(0.31, 3.05) | 2.31(0.99, 5.36) | 3.75(2.03, 6.90) |  |
| Unemployed (Ref) | 1 | 1 | 1 |  |
| **Menstrual cycle** |  |  |  | *0.335* |
| Irregular | 0.64(0.08, 5.01) | 0.27(0.04, 2.02) | 1.5(0.77, 2.94) |  |
| Regular | 1 | 1 | 1 |  |
| **Menstrual status\*** |  |  |  | *<0.001* |
| Menopause | 2.19(0.58, 8.26) | 2.29(0.94, 5.62) | 4.32(2.50, 7.45) |  |
| Pre-menopause (Ref) | 1 | 1 | 1 |  |
| **Parity\*** |  |  |  | *<0.001* |
| Nulliparous | 0.43(0.13, 1.45) | 0.15(0.05, 0.44) | 0.10(0.04, 0.22) |  |
| Parous (Ref) | 1 | 1 | 1 |  |
| **Contraception use\*** | 6.34(1.38, 29.20) | 10.14(3.02, 34.08) | 3.52(2.00, 6.19) | *<0.001* |
| **Family history of BC\*** |  |  |  | *0.069* |
| Yes | 4.52(0.94, 21.84) | 2.35x10<sup>-6</sup>(-, -) | 2.60(1.07, 6.30) |  |
| No (Ref) | 1 | 1 | 1 |  |
| **Smoking history** |  |  |  | *0.532* |
| Yes | 3.67x10<sup>-6</sup>(-, -) | 3.67x10<sup>-6</sup>(-, -) | 2.74(0.72, 10.37) |  |
| No (Ref) | 1 | 1 | 1 |  |
| **Alcohol history** |  |  |  | *0.953* |
| Yes | 1.18x10<sup>-6</sup>(-, -) | 0.56(0.07, 4.24) | 0.91(0.31, 2.64) |  |
| No (Ref) | 1 | 1 | 1 |  |
| **Exercises\*** |  |  |  | *<0.001* |
| Yes | 0.86(0.27, 2.71) | 0.36(0.16, 0.84) | 0.36(0.21, 0.62) |  |
| No (Ref) | 1 | 1 | 1 |  |

Key: Reference category for outcome is Benign, \* Variables eligible for inclusion in the multivariable model (p≤0.05), cOR; crude odds ratio, CI; confidence interval.
https://doi.org/10.7910/DVN/2OXWTA

#### Multivariable multinomial analysis

Based on results from the multivariable analysis, parity, contraceptive use, and exercise were found to be significant predictors of breast lesions at less than 5% significance level. Table 4 illustrates the odds ratios and confidence intervals for the breast lesion predictors across the three categories of breast lesion outcomes (atypical, suspicious, and malignant lesions).

**Table 4:** Multivariable analysis of the risk factors for breast lesions among women with breast lumps attending two select teaching and referral health facilities in Kenya using a multinomial logistic regression model.

| Factors | Values | Atypical lesions<br>aOR (95% C.I) | Suspicious of malignancy<br>aOR (95% C.I) | Malignant lesions (Breast cancer)<br>aOR (95% C.I) | *p-value* |
| --- | --- | --- | --- | --- | --- |
| Parity | Nulliparous | 0.79(0.21, 2.9) | 0.23(0.07, 0.71) | 0.11(0.05, 0.26) | 0.001 |
|  | Parous | 1 | 1 | 1 |  |
| Exercises | Yes | 0.92(0.28, 2.98) | 0.33(0.14, 0.78) | 0.31(0.18, 0.55) | 0.001 |
|  | No | 1 | 1 | 1 |  |
| Contraception | Yes | 5.73(1.14, 28.94) | 6.02(1.73, 20.95) | 1.82(0.99, 3.34) | 0.002 |
|  | No | 1 | 1 | 1 |  |

Key: aOR; adjusted odds ratio
https://doi.org/10.7910/DVN/2OXWTA

The odds of atypical, suspicious, and malignant diagnoses in nulliparous women are about one-tenth, close to a quarter, and four-fifths respectively [aOR: 0.11; 95% CI: 0.05-0.26; p=0.001, aOR: 0.23; 95% CI: 0.07-0.71, p=0.001; aOR: 0.79; 95% CI: 0.21-2.9, p=0.001] controlling for exercise and contraceptive use.

The odds of a diagnosis of atypical lesions is slightly less than a third (aOR: 0.31; 95% CI: 0.18-0.55, p=0.001), for suspicious lesions is a third (aOR: 0.33; 95% CI: 0.14-0.78, p=0.001) and for malignant lesions is more than three quarters (aOR: 0.92; 95% CI: 0.28-2.98, p=0.001), in women who do regular exercise controlling for parity and contraceptive use.

The odds of diagnosing atypical lesions in women who use contraceptives are more than five times (aOR: 5.73; 95% CI: 1.14-28.94, p=0.002), for suspicious lesions is more than six times (aOR: 6.02: 95% CI: 1.73-20.95, p=0.002) while for malignant lesions it is more than one (aOR: 1.8; 95% CI: 0.99-3.34, p=0.002) controlling for parity and exercises.

## Discussion

The key findings of this study suggest that parity, exercise, and contraceptive use are significant predictors of breast cancer and other breast lesions among women in Kenya. Nulliparity was found to have a substantial protective effect against the development of both suspicious lesions of the breast and BC. This is a contradictory finding to other studies in the literature that identified parity as protective in the development of breast cancer, especially those that are ER+/PR+ [18-20]. However, our findings correlate with some studies that have reported parous women to be more likely to develop triple-negative breast cancer [21-24]. This means that parity is not protective against all types of BC as conventionally documented. Although nulliparous women had lower odds of developing atypical lesions, the difference was not statistically significant. Nevertheless, there is an indication that the likelihood of developing atypical breast lesions is lower in women who have never given birth.

In this study, regular exercise was found to be significantly associated with a lower likelihood of BC and other breast lesions. Comparing respondents who didn’t regularly exercise, participants who exercised were less likely to have BC and less likely to be diagnosed with suspicious lesions. There is strong evidence to support that physical exercise has a significant impact on reducing the odds of developing BC [25, 26]. This is mainly because exercise modulates BMI and reduces obesity, which are both risk factors for BC [27]. In addition, physical activity is associated with a significantly delayed onset of BC among breast cancer gene 1 and breast cancer gene 2 (BRCA1/2) mutation carriers [28, 29]. This is because exercise is known to enhance the body’s Deoxyribonucleic acid (DNA) repair mechanisms, potentially correcting mutations that could lead to cancer. Though the odds of developing atypical breast lesions in this population were lower, the difference between those who reported having regular exercise and those who had never had regular exercise was not statistically different.

Our study established that participant who used hormonal contraceptives had higher odds of being diagnosed with BC. These findings compare with those of a study done in Jordan which indicated that regular use of oral contraception [OCs] increased the risk of BC [30]. Contraception use had also been associated with a substantially increased risk of developing suspicious lesions among contraception users and atypical lesions in this study.

In contraceptive usage, almost an equal number of participants had used compared to those who had not. A no-method preference has been reported as the second most preferred in some studies [31]. Our findings compare with data from 47 countries which indicated that 40.9% of women in need of contraception were not using any methods. Moreira and others have explored reasons for the non-use of contraception despite the unmet need. Major reasons for non-use included, health concerns, infrequent sex, and opposition from others [32]. Religious reasons and cultural factors may be the other reasons for non-use in our study population.

In terms of the breakdown of various contraceptives and their preference among women, the oral contraceptive was preferred. This finding is consistent with those reported in the literature [33]. Our study also indicates multiple uses of hormonal contraception over time, where the same individuals had used oral, injection, or implant. Similar findings have been reported previously in the US [34]. Simultaneous use of more than one contraception during sex has also been reported in the literature [35].

Patients’ demographic and socioeconomic characteristics are important considerations in BC diagnosis and management. Most participants in this study had a minimum of secondary education suggesting a level of breast health awareness and positive health-seeking behavior in this population. Most participants were of the Kikuyu ethnic community. This is likely since this is the second most populous ethnic group in Nakuru County and the largest ethnic group in Kenya. In addition, Nairobi City is in close proximity to Central Kenya, a geographical region historically occupied by Kikuyus. Kikuyus being the majority attending the Breast FNA Clinic in KNH could be purely due to the case of accessibility in this case rather than other factors.

Concerning parity, a small difference was noted between the parous and nulliparous BC patients. Ideally, since many of the women in this study were in their thirties, one would have expected most of them to be parous, but this was not the case. This could have been due to lifestyle and priority changes. Most ladies would want to focus on their career paths before they start getting children while others plan to have babies after the age of 30 [36].

The prevalence of other known risk factors for cancer including family history, alcohol use, and smoking were very low among our study participants. Generally, a proportion of cancer cases are attributable to smoking and high alcohol consumption [37]. Exercises or no exercise/lack of it was almost equally practiced among our study participants. This is important to note since our participants seemed to do exercises at a higher percentage [about 50%] than one would expect in the general population. Data from Europe indicate that 60% of individuals aged 15 and above rarely or never participate in exercise or sports [38]. While physical exercise contributes to a healthy life, most people do not engage in exercise. Physical inactivity is a major public health problem for people of all ages and is currently rated as the fourth-highest global risk factor for mortality and pathologies such as type 2 diabetes, coronary heart disease, and certain types of cancer [39]. Study participants in the current study probably participate in exercise due to increased awareness and need, based on their medical condition, or it is part of their lifestyle since an equal percentage did not as well.

Overall, from the univariable analysis, the factors that were found to be significantly associated with breast lesions included age, ethnicity, level of education, employment status, family history of BC, and exercise. The findings from this study correlate with those of other studies. The risk of BC increases with age [40]. Incidence also increases and reaches its peak in the age of menopause and then gradually decreases or remains constant [41]. Several studies have reported racial and ethnic variation in BC where a minority of women present with higher-stage BC than white women [42, 43]. Such differences in race and ethnicity have also been discussed by others [42, 44, 45].

Education level and employment status were significantly associated with breast cancer in this study. This is consistent with other studies [46, 47]. This could be mainly because employment and education are associated with awareness, quality of life, and exposure to other risk factors like alcohol and smoking, as well as postponement of childbearing and number of children while women pursue further education and careers [48]. Family history is an important risk factor for BC as demonstrated in our study. This important association has been demonstrated previously and in other studies [49-53].

## Conclusion

From this study, exercise, parity, and hormonal contraception were found to be significant predictors of breast cancer and other breast lesions. These findings warrant consideration of BC risk reduction through empowering women to make informed reproductive choices and the choice of contraception use and weight management.

## Data Availability

The data underlying the results presented in the study are available in Harvard Dataverse. https://doi.org/10.7910/DVN/2OXWTA.

## Acknowledgement

The authors wish to express their sincere gratitude to the study participants for contributing to the research. They also acknowledge the University of Nairobi s Building Capacity for Writing Scientific Manuscripts (UANDISHI) Program at the Faculty of Health Sciences for the training on manuscript writing.

## Author Contributions

Conceptualization: Josephine Nyabeta Rioki, Emily Rogena.

Data curation: Josephine Nyabeta Rioki, Marshal Mweu.

Formal analysis: Josephine Nyabeta Rioki, Marshal Mweu.

Funding acquisition: Josephine Nyabeta Rioki, Elijah Songok.

Investigation: Josephine Nyabeta Rioki, Lucy Muchiri, Marshal Mweu, Elijah Songok, Emily Rogena.

Methodology: Josephine Nyabeta Rioki, Marshal Mweu.

Project administration: Josephine Nyabeta Rioki, Lucy Muchiri, Marshal Mweu, Elijah Songok, Emily Rogena.

Supervision: Lucy Muchiri, Marshal Mweu, Elijah Songok, Emily Rogena.

Writing – original draft: Josephine Nyabeta Rioki, Joseph Mwangi.

Writing – review & editing: Josephine Nyabeta Rioki, Lucy Muchiri, Marshal Mweu, Elijah Songok, Emily Rogena.

## Supporting information

**S1 File:** Harvard Dataverse: Replication data for: Factors associated with breast lesions among women attending select teaching and referral health facilities in Kenya: a cross-sectional study, https://doi.org/10.7910/DVN/2OXWTA.

